# Examining Unit Costs for COVID-19 Case Management in Kenya

**DOI:** 10.1101/2020.10.08.20209684

**Authors:** Edwine Barasa, Angela Kairu, Wangari Nganga, Marybeth Maritim, Vincent Were, Samuel Akech, Mercy Mwangangi

## Abstract

**Introduction:** Case management for COVID-19 patients is one of key interventions in country responses to the pandemic. Countries need information on the costs of case management to inform resource mobilization, planning and budgeting, purchasing arrangements, and assessments of the cost-effectiveness of interventions. We estimated unit costs for COVID-19 case management for patients with asymptomatic, mild to moderate, severe, and critical COVID-19 disease in Kenya.

**Methods:** We estimated per patient per day unit costs of COVID-19 case management for patients that are asymptomatic and those that have mild to moderate, severe, and critical symptoms. For asymptomatic and mild to moderate patients, we estimated unit costs for home-based care and institutional (hospitals and isolation centers). We used an ingredients approach, adopted a health system perspective and patient episode of care as our time horizon. We obtained data on inputs and their quantities from COVID-19 case management guidelines, home based care guidelines, and human resource guidelines, and augmented this with data provided by three public covid-19 treatment hospitals in Kenya. We obtained input prices for services from a recent costing survey of 20 hospitals in Kenya and for pharmaceuticals, non-pharmaceuticals, devices and equipment from market price databases for Kenya.

**Results:** Per day per patient unit cost for asymptomatic patients and patients with mild to moderate COVID-19 disease under home based care are KES 1,993.01 (USD 18.89) and 1995.17 (USD 18.991) respectively. When these patients are managed in an isolation center of hospital, the same unit costs for asymptomatic patients and patients with mild to moderate disease are 7,415.28 (USD 70.29) and 7,417.44 (USD 70.31) respectively. Per day unit costs for patients with severe COVID-19 disease managed in general hospital wards and those with critical COVID-19 disease admitted in intensive care units are 12,570.75 (USD 119.16) and 59,369.42 (USD 562.79).

**Conclusion:** COVID-19 case management costs are substantial. Unit costs for asymptomatic and mild to moderate COVID-19 patients in home-based care is 4-fold lower compared institutional care of the same patients. Kenya will not only need to mobilize substantial resources to finance COVID-19 case management but also explore additional service delivery adaptations that will reduce unit costs.

## Background

The World Health Organization, declared COVID-19, caused by the severe acute respiratory syndrome coronavirus 2 (SARS-CoV-2), a pandemic on March 11 2020 (1). The pandemic has spread to almost all countries and territories worldwide, infecting millions of individuals and causing many deaths (2). To effectively respond to the pandemic, countries need to identify cost-effective interventions, plan and mobilize resources to deploy these interventions in ways that enhance health system goals that include equitable access, efficiency, quality, and financial risk protection. To effectively achieve this, countries need information on the unit costs of these interventions.

One such area of intervention is case management of patients that test positive for COVID-19. While most people with COVID-19 are either asymptomatic or develop only mild to moderate disease, estimates from Asia, Europe and the US show that approximately 15% develop severe disease that requires oxygen support and management in general hospital wards, and 5% have critical disease, likely to require mechanical ventilation and may develop complications such as respiratory failure, acute respiratory distress syndrome (ARDS), thromboembolism, sepsis and septic shock, and multi-organ failure such as cardiac and acute kidney injury (3–5). Patients with critical disease require intensive care unit care (4).

Information about the unit costs for COVID-19 case management is useful in mobilizing resources and planning and budgeting for this intervention by policy makers. It is also useful in formulating appropriate healthcare purchasing arrangements by informing the development of provider payment mechanisms and rates, and as estimates to parametize cost-effectiveness models to assess the value for money of COVID-19 interventions. In this paper, we present an analysis to estimate the unit costs for COVID-19 case management in Kenya. Kenya has developed case management guidelines for COVID-19 and is pondering appropriate financing mechanisms and purchasing arrangements for case management as part of its health system response to the pandemic. Evidence of case management unit costs will therefore find utility in informing these policy decisions and could be adapted and adjusted to facilitate application in other countries with similar contexts.

## Methods

### Intervention description

We costed case management for COVID-19 for four clinical severities as defined by the Kenya ministry of health COVID-19 case management guidelines (6). These are:

- Asymptomatic COVID-19 patients
- Covid-19 patients with mild to moderate symptoms
- Covid-19 patients with severe symptoms
- COVID-19 patients in critical condition

Kenya introduced a policy for home-based care for COVID-19 patients that are asymptomatic or have mild to moderate symptoms (7). This policy is intended to reduce the burden of case management by hospitals and isolation centers and reduce the costs of management. Patient eligibility criteria for home-based care are a) laboratory confirmed COVID-19 b) asymptomatic patients or patients with mild to moderate symptoms of COVID-19 c) absence of co-morbidities, d) access to a suitable space for home-based isolation (7). The decision to adopt home-based care for a patient is made after an assessment of the suitability of the patients’ home environment by a trained healthcare worker. Patients on home-based care are required to self-isolate at home and self-report symptoms daily to a healthcare provider using a mobile phone application (7). The patients are only transferred to a hospital if they develop severe symptoms. Patients that do not qualify for home-based care receive institutional care – in a hospital or an isolation center. We therefore costed case management for asymptomatic patients and patients with mild to moderate symptoms for both scenarios – home-based care and institutional care.

According to the Kenya covid-19 case-management guidelines, patients with severe symptoms are admitted for inpatient general hospital care with the option of oxygen therapy when needed (6). Patients in critical condition are admitted to the intensive care unit (ICU) with the option for mechanical ventilation when needed (6).

### Costing approach, perspective, and time horizon

We used an ingredients approach to costing (8), which entails the identification of relevant inputs that are used to deliver COVID-19 case management, their quantities, and their monetary value. We costed the case management of COVID-19 patients from a health system perspective and used one patient episode of care as our time horizon.

### Type of costs and unit costs estimated

We estimated economic costs which include the monetary value of inputs whether they are accompanied with financial outlays or not. We estimated the following unit costs for a) Per patient completing treatment cost of COVID-19 case-management, and b) Per patient, per day cost of COVID-19 case management for the following patient categories and delivery strategies:

1. Asymptomatic COVID-19 patients on home-based care
2. Asymptomatic COVID-19 patients on institutional care (admitted in hospitals or isolation centers)
3. Mild to moderate symptomatic COVID-19 patients on home-based care
4. Mild to moderate symptomatic COVID-19 patients on institutional care (admitted in hospitals or isolation centers)
5. COVID-19 patients with severe symptoms admitted in hospitals
6. COVID-19 patients with critical disease admitted in intensive care units

#### Measuring resource use

For each of the unit costs, we costed all the direct and ancillary inputs that go into the delivery of the case management. Broadly, this included health facility “hotel costs” which included non-clinical costs of inpatient accommodation and overheads (e.g. management, electricity, water, infrastructure), staff time, pharmaceutical and non-pharmaceuticals patient level interventions, laboratory test, radiology costs, and costs for personal protective equipment (figure 1).

**Figure 1:**
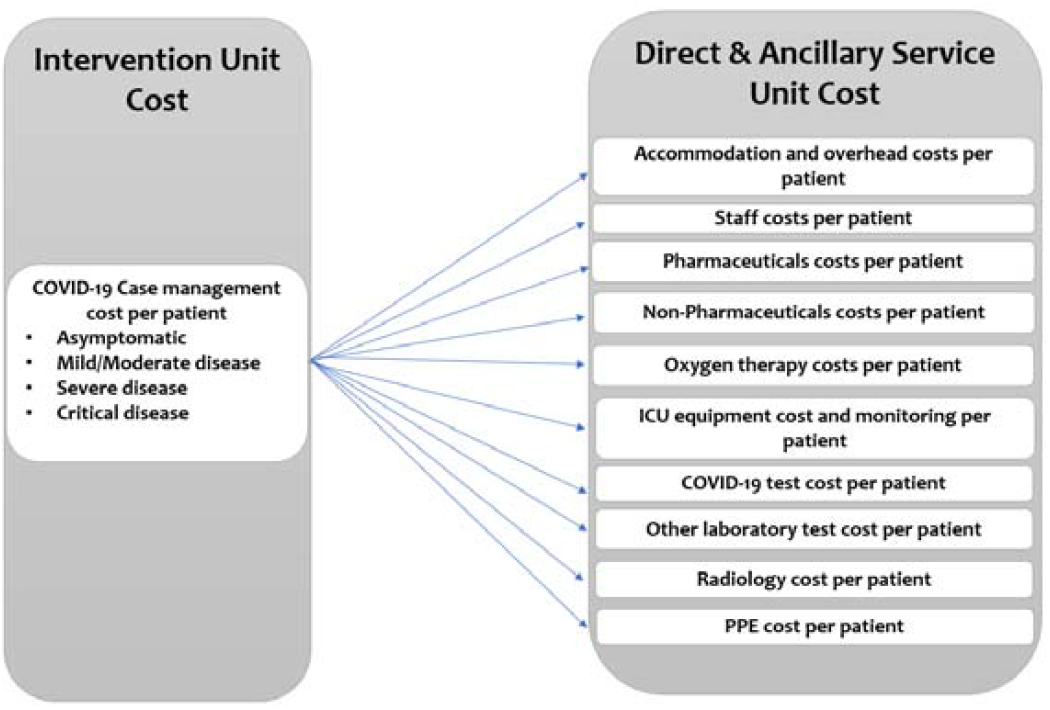
COVID-19 Case Management Unit Costs

We obtained data on inputs and their quantities from billing reports from 3 public health facilities designated as COVID-19 treatment centers, and the Kenya COVID-19 guidelines on a) case management (6), b) home-based care (7), and c) human resources requirements (9). These guidelines outlined the interventions, resources, and resource quantities that ought to be used for case management of COVID-19 patients in Kenya. We obtained data on input costs from recent primary cost data whose estimates were appropriate to the COVID-19 pattern of care, primary data from 3 COVID-19 public treatment healthcare centers, and market prices.

### Accommodation and overheads

We obtained per day costs for accommodation and overheads from median costs reported in a cost analysis of 20 healthcare facilities in Kenya in 2018. Accommodation and overhead costs include costs for non-medical inputs including accommodation, management, catering, laundry and cleaning, electricity and water.

### Staffing

We obtained data on the type of staff, and time spent on patients from the Kenya ministry of health human resource guidelines for the management of COVID-19 and from reports of actual staff allocation in 3 public COVID-19 treatment centers (supplementary file 1). We obtained staff costs from official public sector staff salaries data provided to us by county governments in Kenya.

### Pharmaceuticals and Non-pharmaceutical (fluids, devices etc), and personal protective equipment

Supplementary file 1 outlines the list of medicines and non-pharmaceuticals that are used in the management of COVID-19 in Kenya. These were obtained from COVID-19 clinical guidelines for Kenya and billing information from 3 public COVID-19 treatment centers. We obtained costs for medical equipment from market prices for local suppliers, and medicines, and non-pharmaceuticals from market prices reported in the 2020 Kenya drug index catalogue (10). This catalogue provides current prices of all available brands of pharmaceuticals and non-pharmaceuticals in Kenya. We used median costs of available generic medicines or brands of non-pharmaceuticals. We only used the price of originator medicines when they were the only ones available in the Kenyan market (i.e. there was no generic alternative). Costs for personal protective equipment were from market prices.

### Laboratory and Radiology costs

Supplementary file 1 outlines the laboratory and radiology test carried out on COVID-19 patients in Kenya. These were obtained from COVID-19 clinical guidelines for Kenya and billing information from 3 public COVID-19 treatment centers. We obtained information on inputs, their quantities and costs for the PCR COVID-19 test from one of the national COVID-19 testing centers in Kenya. We used fees charged for laboratory tests and radiology by health facilities from the 2018 cost survey of 20 healthcare facilities in Kenya to represent their costs.

### Oxygen and Equipment costs

We obtained oxygen and equipment costs from local market prices. Medical equipment was assumed to have a useful life of 5 years.

#### Transferring costs over time

We used gross domestic product (GDP) deflators for Kenya to adjust 2018 input costs to 2020 and used an exchange rate of 1 United States Dollar (USD)=105.49 Kenya Shillings (KES), derived from oanda.com and accessed on 30^th^ June 2020, to convert Kenya shillings to USD. We report our findings in 2020 KES and USD.

## Results

### Unit costs for COVID-19 patients that are asymptomatic and mild to moderate disease

Table 1 and 2 outlines the unit costs for COVID-19 case management of patients that are asymptomatic and those that have mild to moderate symptoms respectively. Per day unit costs for home-based care case management of asymptomatic patients and patients will mild to moderate symptoms is KES 1,993.01 (USD 18.89) and 1995.17 (USD 18.991) respectively. When these patients are managed in a general hospital ward or an isolation center, per day unit costs for asymptomatic patients and patients with mild to moderate symptoms are estimated to be 7,415.28 (USD 70.29) and 7,417.44 (USD 70.31) respectively. There is a negligible cost difference in the management of asymptomatic patients and patients with mild to moderate disease. This is because patients with mild to moderate disease only incur additional costs for symptom relievers such as paracetamol. PPE accounts for the largest share of costs for asymptomatic and mild to moderate disease. This is because these patients receive minimal interventions. They for instance hardly receive any medicines, do not undergo any radiological tests, and do not receive supplemental oxygen.

**Table 1:**
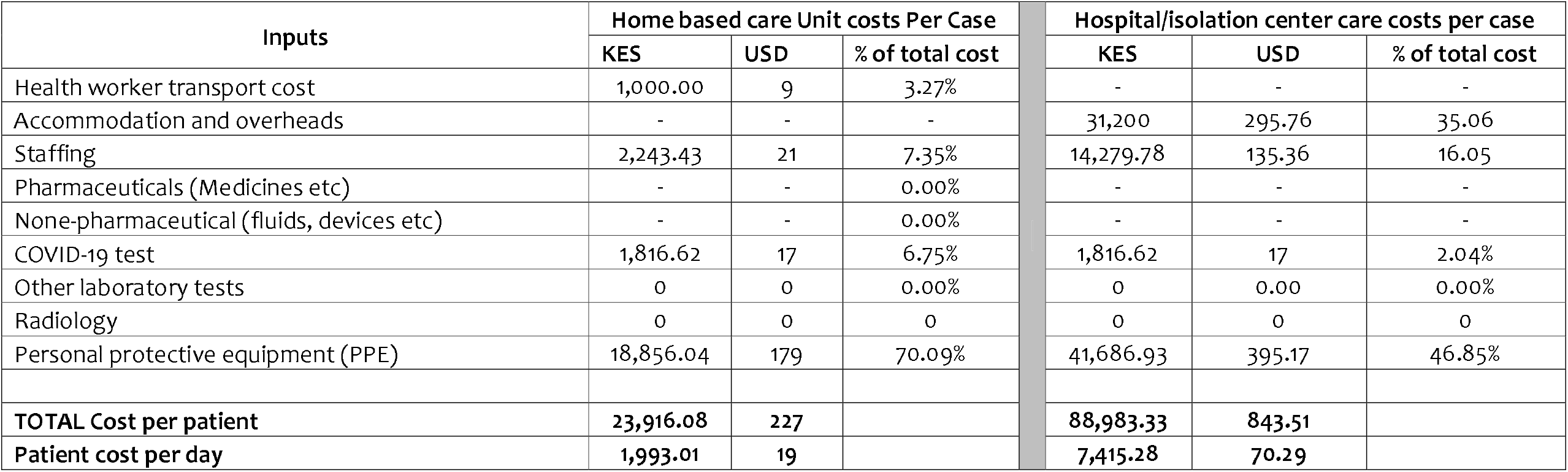
COVID-19 Case management unit costs for asymptomatic patients

| Inputs | Home based care Unit costs Per Case |  |  | Hospital/isolation center care costs per case |  |  |
| --- | --- | --- | --- | --- | --- | --- |
|  | KES | USD | % of total cost | KES | USD | % of total cost |
| Health worker transport cost | 1,000.00 | 9 | 3.27% | - | - | - |
| Accommodation and overheads | - | - | - | 31,200 | 295.76 | 35.06 |
| Staffing | 2,243.43 | 21 | 7.35% | 14,279.78 | 135.36 | 16.05 |
| Pharmaceuticals (Medicines etc) | - | - | 0.00% | - | - | - |
| None-pharmaceutical (fluids, devices etc) | - | - | 0.00% | - | - | - |
| COVID-19 test | 1,816.62 | 17 | 6.75% | 1,816.62 | 17 | 2.04% |
| Other laboratory tests | 0 | 0 | 0.00% | 0 | 0.00 | 0.00% |
| Radiology | 0 | 0 | 0 | 0 | 0 | 0 |
| Personal protective equipment (PPE) | 18,856.04 | 179 | 70.09% | 41,686.93 | 395.17 | 46.85% |
| **TOTAL Cost per patient** | **23,916.08** | **227** |  | **88,983.33** | **843.51** |  |
| **Patient cost per day** | **1,993.01** | **19** |  | **7,415.28** | **70.29** |  |

**Table 2:**
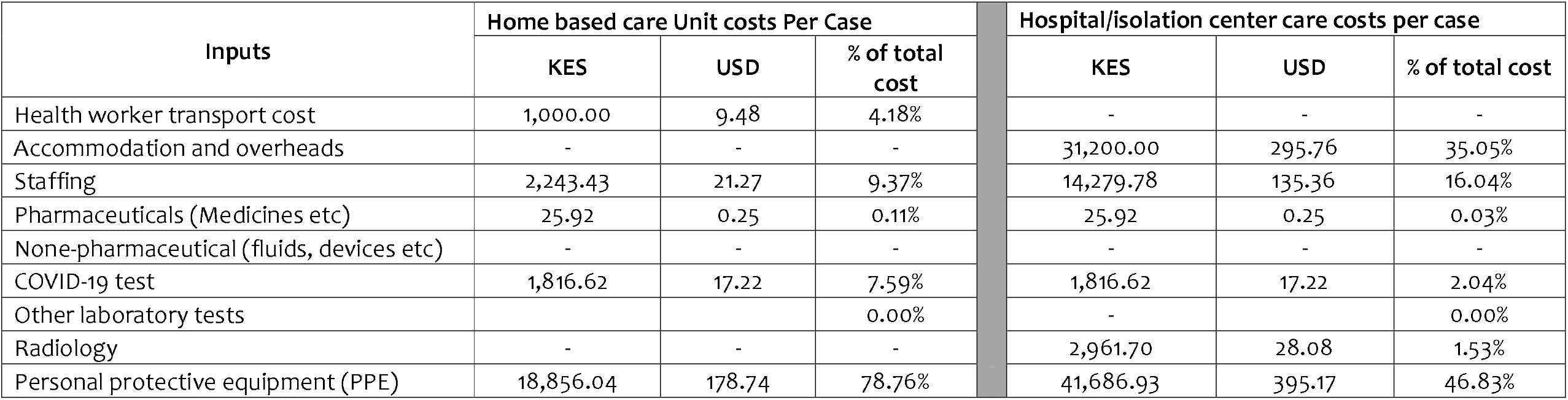
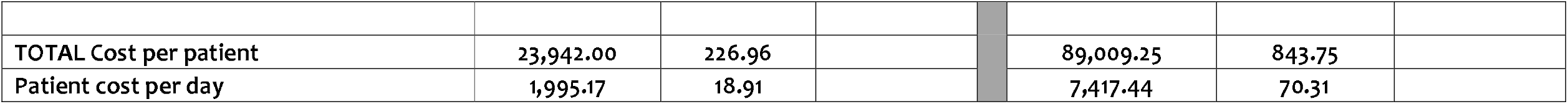
COVID-19 Case management unit costs for patients with mild to moderate symptoms

Unit costs under home-based care are substantially lower than unit costs for institutional care (hospitals or isolation centers). This is because home-based care avoids hospital accommodation and overhead costs and is characterized by minimal health worker-patient physical interactions and hence saves on staffing, accommodation, and personal protective equipment costs. The only cost associated with health worker interactions are transport costs to assess the patient’s home for suitability of home - based care.

### Patients with severe and critical COVID-19 disease

Table 3 outlines the unit cost for case management of COVID-19 patients with severe and critical disease. Per day unit costs for case management for severe disease are estimated to be KES 12,570.75 (USD 119.16). Per day unit costs for case management for critical disease are estimated to be KES 59,369.42 (USD 562.79). Severe disease patient costs differ from those of hospitalized patients with mild to moderate disease because of additional costs for pharmaceuticals (e.g. antibiotics), staff, and oxygen therapy. Patients with critical disease incur higher intensive care related costs that include specialist staff (e.g. critical care physicians, anaesthetists) and more staff time per patient, pharmaceuticals (antibiotics and anaesthesia medicine) and non-pharmaceuticals (e.g. total parenteral nutrition), mechanical ventilation, other monitoring equipment costs. Pharmaceuticals and PPE are the key contributors of costs for the management of patients with severe COVDI-19. This is because more health workers are involved in the care of these patients (increasing PPE costs) and the patients receive pharmaceutical and non-pharmaceutical interventions such as antibiotics, fluids and oxygen. Staff costs contribute the largest share of costs for critical COVID-19 patients because these patients not only need more numbers of staff, they also need more staff time (health worker patient ratio) and more specialised care (such as physicians and anaesthetists) which cost more.

**Table 3:** COVID-19 Case management unit costs for patients with severe and critical COVID-19 disease

| Inputs | Unit costs for Severe COVID-19 disease |  |  | Unit costs for critical COVID-19 disease |  |  |
| --- | --- | --- | --- | --- | --- | --- |
|  | KES | USD | % of total cost | KES | USD | % of total cost |
| Accommodation and overheads | 31,200.00 | 295.76 | 20.68% | 48,000.00 | 455.01 | 6.74% |
| Staffing | 19,835.18 | 188.03 | 13.15% | 350,512.56 | 3,322.65 | 49.20% |
| Pharmaceuticals (Medicines etc) | 55,224.45 | 523.49 | 36.61% | 71,946.61 | 682.01 | 10.10% |
| None-pharmaceutical (fluids, devices etc) | 2487.2 | 23.58 | 1.65% | 4,527.20 | 42.92 | 0.64% |
| COVID-19 test | 1,816.62 | 17.22 | 1.20% | 1,816.62 | 17.22 | 0.25% |
| Other laboratory tests | 10,817.77 | 102.55 | 7.17% | 21,817.77 | 206.82 | 3.06% |
| Radiology | 2,961.70 | 28.08 | 1.96% | 2,961.70 | 28.08 | 0.42% |
| Personal protective equipment (PPE) | 44,292.37 | 419.86 | 29.36% | 243,173.78 | 2,305.14 | 34.13% |
| Oxygen therapy | 13,413.79 | 127.15 | 8.89% | 15,676.79 | 148.61 | 2.20% |
| Equipment costs (including ventilator) and monitoring in ICU | - | - | - | 12,887.60 | 122.17 | 1.81% |
| **TOTAL Cost per patient** | **150,849.06** | **1,429.96** |  | **712,433.01** | **6,753.43** |  |
| **Patient cost per day** | **12,570.75** | **119.16** |  | **59,369.42** | **562.79** |  |

## Discussion

This study presents evidence on the costs of COVID-19 case management in Kenya. Specifically, its presents unit costs for the management of asymptomatic patients, and COVID-19 patients with mild to moderate, severe and critical disease. For asymptomatic and mild to moderate disease patients, we analyse costs for home-based care and those for care in isolation centers or general hospital wards. The findings show that COVID-19 case management costs are substantial for all treatment categories and increase as severity increases. These high case management costs have several implications. First, these costs will put a fiscal strain to LMIC health system’s like Kenya because of existing resource challenges. Kenya will need to actively mobilize both domestic and donor resources to meet these costs. Second, Kenya, and other LMIC may need to adapt case management guidelines further to improve efficiencies and affordability without compromising quality of care. A good example is the home-based care strategy that Kenya has already adopted for patients that are asymptomatic and those with mild to moderate disease. Our findings show that unit costs for home-based care are 4 times lower than those for institutional care resulting in substantial costs savings. However, not all asymptomatic and mild to moderate disease patients qualify for home-based care, and some will still need to be institutionalized because they are high risk (e.g. have co-morbidities) or their home environments are unsuitable for home-based care. These include individuals living in low income housing including urban informal settlements. These patients will still need to be institutionalized. However, the use of the same self-reporting mobile technology could for instance minimize health worker patient interactions and substantially reduce both staffing and PPE costs even for these hospitalized patients. Further, a lower cadre of health workers could also be used to monitor patients in isolation centers. Other adaptations could target cost drivers such as length of stay and discharge protocols, with patients discharged to home-based care as soon as their symptoms improve from severe to mild/moderate. A third implication is that Kenya and other LMICs will need mechanisms to protect COVID-19 patients from the financial burden of healthcare costs to access COVID-19 services. If these costs are passed to patients as direct healthcare costs, they will result in substantial levels of catastrophic healthcare expenditures and impoverishment. There is therefore an urgent need for Kenya and LMICs in similar situations to develop a prepayment mechanism to provide financial risk protection to patients and households against the financial hardship that they will face if required to pay for COVID-19 case management costs out of pocket.

This analysis has several limitations. First, we extensively relied on normative guidelines assumptions and cost data from previous studies, and only collected data from 3 COVID-19 treatment centers because it was impossible to carry out real world extensive data collection given existing physical distancing restrictions. However, this limitation is mitigated by the fact that there is no specific COVID-19 treatment and that COVID-19 case management reflects management of non-COVID-19 patients with the same symptoms. The cost survey we relied on was also relatively recent. Second, this analysis presents data for public sector costs. Private sector costs would be useful given that the COVID-19 response will require governments to purchase services from both the public and private sector in settings like Kenya where the private sector plays a significant role in healthcare service provision. Third, input costs for some items, especially PPE’s are volatile because of market disruptions and are likely to stabilize much later, and thus reducing the unit costs of case management. We have however used market prices 8 months into the pandemic which are likely to be closer to the stable prices in the future rather than the costs at the beginning of the pandemic. Third, we did not analyse costs for the range of possible COVID-19 complications. This is because unit costs exist for some of these, for instance for kidney replacement therapy, and costing the entire range of possible complications would require data what were not available given the fieldwork restrictions as a result of physical distancing measures. These limitations not-withstanding, the estimates we present will be useful in informing Kenya’s resource mobilization for the COVID-19, budgeting and planning, as well as informing the country’s plan to develop appropriate purchasing mechanisms that include provider payment mechanism and rates that are appropriate for COVID-19. The estimates will also find utility in parametizing cost-effectiveness models for COVID-19 interventions as and when they become available such as a COVID-19 vaccine. While these costs estimates have been developed for Kenya, they could potentially find use and applicability in other LMIC’s with comparable settings after adapting and adjusting with country specific assumptions.

## Data Availability

All the data used in this study is included in the manuscript and supplementary files

