## Supplementary file 1 for "Examining Unit Costs for COVID-19 Case Management in Kenya"

**Table A1: Staff providing direct* care to COVID-19 patients**

| **Staff** | **Asymptomatic & mild to moderate COVID patients (home based care)** | **Asymptomatic & mild to moderate COVID patients (home based care)** | **Severe COVID patients** | **Critical COVID patients** |
| --- | --- | --- | --- | --- |
| Physician/ Medical doctor | - | √ | √ | √ |
| Clinical officer | √ | - | - | - |
| Community health worker | √ | - |  | √ |
| Nurse | - | √ | √ | √ |
| Physiotherapist | - | - | √ | √ |
| Anaesthetist | - | - | - | √ |
| Nutritionist | - | - | - | √ |

*Staff providing indirect care are costed as part of specific services e.g. laboratory staff, pharmacy staff, cleaners, porters etc

**Table A2: Medicines used on COVID-19 patient care**

| **Medicine** | **Asymptomatic COVID-19 patients** | **Mild COVID-19 patients** | **Severe COVID patients** | **Critical COVID patients** |
| --- | --- | --- | --- | --- |
| Paracetamol | - | √ | √ | √ |
| Amoxicillin+clavulanic acid | - | - | √ | √ |
| Azithromycin | - | - | √ | √ |
| Subcutaneous enoxaparin | - | - | √ | √ |
| Dexamethasone | - | - | √ | √ |
| **Medicines used for intubation and sedation** |  |  |  |  |
| Ketamine | - | - | - | √ |
| Propofol | - | - | - | √ |
| Succycholine | - | - | - | √ |
| Rocuronium | - | - | - | √ |
| Morphine | - | - | - | √ |
| Ketamine | - | - | - | √ |

**Table A3: Laboratory and radiological tests carried out on COVID-19 patients**

| **Test** | **Frequency per treatment episode** | **Asymptomatic COVID-19 patients** | **Mild to moderate COVID-19 patients** | **Severe COVID patients** | **Critical COVID patients** |
| --- | --- | --- | --- | --- | --- |
| **COVID-19 test** | 1 | √ | √ | √ | √ |
| **Other laboratory tests** |  |  |  |  |  |
| Full hemogram | 1 | - | - | √ | √ |
| UECs | 1 | - | - | √ | √ |
| LFTs | 1 | - | - | √ | √ |
| Random blood sugar | Daily | - | - | √ | √ |
| C-reactive protein levels | 1 | - | - | √ | √ |
| D-dimers | 1 | - | - | √ | √ |
| Ferritin levels | 1 | - | - | √ | √ |
| Rapid HIV test | 1 | - | - | √ | √ |
| Blood gas analysis (BGA) | Daily | - | - | - | √ |
| **Radiology** |  |  |  |  |  |
| Chest X-ray (film) | 1 | - | - | √ | √ |

**Table A4: PPE’s used by a healthcare workers and other staff in a COVID-19 treatment facility**

| **PPE** | **Useful life** | **Quantity used by a healthcare worker per shift** |
| --- | --- | --- |
| N-95 Masks | 1 | 4 |
| Tyvek Suits | 1 | 1 |
| Surgical Gowns | 1 | 1 |
| Nitrile Gloves | 1 | 10 |
| Latex Gloves | 1 | 10 |
| Disposable Head | 1 | 4 |
| Shoe Covers | 1 | 4 |
| Surgical Masks | 1 | 4 |
| Face Shields | 5 | 1 |
| Goggles | 90 | 1 |
